# Using oral and parenteral formulation of AWaRe antibiotics as a proxy estimate of primary healthcare and inpatient hospital sector use

**DOI:** 10.1101/2025.11.05.25339617

**Authors:** Cherry Lim, Ines Pauwels, Hoa Q Nguyen, Will Cuningham, Mike Thorn, Aislinn Cook, Myo Maung Maung Swe, Ben S Cooper, Benedikt Huttner, Yingfen Hsia, Catrin E Moore, Erika Vlieghe, Koen B. Pouwels, Ann Versporten, Michael Sharland, Global-PPS network

## Abstract

**Background:** Benchmarking antibiotic use across different healthcare sectors is crucial to improve use and implement the UNGA 70% Access target. Many countries only have available aggregate sales data, which do not have sector-specific usage information. The objective of this study is to estimate the proportion of oral antibiotic use across different healthcare sectors.

**Materials/methods:** We used IQVIA MIDAS^®^ Quarterly Sales data and Global Point Prevalence Survey (Global-PPS) inpatient data from eight countries, including Belgium, Canada, China, Netherlands, Philippines, Saudi Arabia, Singapore, and United Kingdom, in 2019. Our analysis focused on Access and Watch antibiotics. In the main analysis, we assumed that all parenteral antibiotics were used exclusively in inpatient settings, an assumption we then relaxed through sensitivity analyses. The observed ratios of oral-to-parenteral antibiotics in the patient-level Global-PPS data were calculated, by dividing the volume of oral antibiotic use by that of parenteral antibiotic use, and then this calculated ratio was multiplied to the IQVIA MIDAS sales data to estimate oral antibiotic use outside of the inpatient sector.

**Results:** The ratios of oral-to-parenteral use among inpatients in the Global-PPS data ranged between 0.05 (95%Credible Interval [CrI]: 0.03-0.09) and 1.01 (95%CrI: 0.56-1.80) in the main analysis. We estimated that overall, less than 7% of national oral antibiotics were used by inpatients, assuming exclusive parenteral use in inpatient settings in the main analysis, and less than 9% when assuming 90% of parenteral use was non-inpatient in the sensitivity analyses.

**Conclusion:** Our results suggest that where patient-level data are unavailable, alternative sources, such as antibiotic import data including routes of administration, can reasonably estimate sector-specific national use.

## Introduction

Antimicrobial resistance (AMR) is a global health threat. It is estimated that the number of deaths due to AMR in 2019 ranged from 1.27 million (95% uncertainty interval [UI]: 0.91-1.71) to 4.95 million (95% UI: 3.62-6.57) globally.^1^ Recognising the important roles of inappropriate antibiotic use in driving the emergence and spread of AMR,^2^ the 79th UN General Assembly set a target for 70% of global human antibiotic use to come from the Access group of WHO’s AWaRe (Access, Watch, Reserve) classification of antibiotics, setting a measurable target for improving antibiotic use.^3^ The development and monitoring of antibiotic use targets at a country level necessitates robust data. Initiatives, such as the Global Point Prevalence Survey of Antimicrobial Consumption and Resistance (Global-PPS),^4^ have contributed to bridging the gap in hospital antibiotic use data, but there is very limited data on primary care antibiotic use to date. The IQVIA MIDAS Quarterly Sales data,^5^ provides useful data on national antibiotic sales trends, but only for 77 countries.^6^ The World Health Organization (WHO) launched the Global Antimicrobial Resistance and Use Surveillance System (GLASS) to support the Global Action Plan to understand antibiotic use and tackle AMR.^7,8^ The WHO GLASS-antimicrobial use (AMU) report on antibiotic data in 2022 suggested oral formulation of antibiotics accounts for over 90% of the total use in most countries.^7^ Most countries only have access to internal import trade data, which provides a broad overview of national antibiotic use, but does not include the healthcare sector-specific use details (i.e. usage in primary-care, inpatients, outpatients, pharmacies and etc) which are needed to inform targeted policy development and implementation.^9^ One approach for collecting community antibiotic use data is to conduct household surveys, but these are costly, time-consuming and therefore not conducted regularly at a country level.^10,11^ Hospital electronic pharmacy databases record longitudinal antibiotic use in the hospital and, when combined with hospital admission data, can be used to analyse aggregated inpatient antibiotic use.^12,13^ However, such databases are challenging to extract or are unavailable in most low and middle-income countries and many high-income countries.

Disparities in data collection methods and sources pose a challenge in combining different data resources for benchmarking usage patterns across inpatient care, outpatient services, and community pharmacies. Moreover, the data gap highlights the need for simple tools to estimate antibiotic use across different healthcare sectors to support policy development, implementation and evaluation of outcomes. The objective of this study was to estimate the proportion of oral antibiotic use across different healthcare sectors. We proposed a new approach of using formulation (oral/parenteral) as a proxy for whether the antibiotic was used in the primary care or hospital sector, when only aggregated antibiotic sales data without sector-specific usage information was available.

## Methods

### Data sources

We obtained antibiotic data from eight countries where data was available from both the IQVIA MIDAS antibiotic sales data and from the Global-PPS antibiotic use data in 2019. The year 2019 was chosen because it was pre-COVID-19. These countries were chosen based on having at least two hospitals in the Global-PPS data collection in 2019. The eight countries from four of the six WHO regions were Belgium, Canada, China, the Netherlands, the Philippines, Saudi Arabia, Singapore, and the United Kingdom. IQVIA collected data on national estimated antibiotic sales, which included antibiotic formulations and sector (“hospital” vs “retail”) from which the data were from, whereas the Global-PPS collected patient-level data on antimicrobial prescribing in hospital facilities.

The hospital antibiotic prescribing data were collected using a 1-day point prevalence survey on inpatient hospital wards according to the 2019 Global-PPS protocol. Participation in the Global-PPS project by hospitals was voluntary, and local ethical approval was obtained where necessary. All data were pseudonymized at the patient level. The Global-PPS data collection methods have been described in detail elsewhere.^4^ For each antibiotic prescription, the prescribed dose on the day of the PPS in DDDs was calculated based on dosing and frequency information and the WHO ATC/DDD list.^14^ The following antibiotics were included: antibacterials for systemic use (ATC J01), nitroimidazole derivatives (ATC P01AB), and antibiotics used as intestinal antiinfectives (ATC A07AA). Since the number of hospitals participating in the PPS varied across countries, the defined daily dose (DDD) was standardised to represent the DDD per 100 surveyed patients per day. Antibiotic prescriptions for children aged under 18 were excluded as currently there is no standard DDD methodology for children.

### Conversion to WHO’s Defined Daily Dose in IQVIA MIDAS antibiotic sales data

We used IQVIA’s New Form Code Classification Guidelines version 2025 to convert antibiotic units and to classify formulations into oral (e.g. tablets, capsules, or suspensions) or parenteral (e.g. ampoules, vials) routes of administration in the IQVIA MIDAS antibiotic sale data. In the original raw data, antibiotic use for a given country and year was presented in kg. DDD is defined as the assumed average maintenance dose per day for adults and was obtained from the ATC/DDD Index website maintained by the Norwegian Institute of Public Health (a WHO Collaborating Centre). We used the kg weight divided by the WHO’s Defined Daily Dose (DDD) to calculate antimicrobial use (AMU) in units of DDD. A minority of antibiotics in the IQVIA MIDAS data did not have a WHO DDD available. For these drugs, we undertook an internet search for recommended daily doses from reputable sources such as National Institute for Health and Care Excellence (NICE) or the manufacturer. By these combined means, we were able to calculate AMU for 97.4% (10,829 out of 11,120 total records of antibiotic regimens) of oral antibiotics and 97.3% (5,421 out of 5,572 total records of antibiotic regimens) of the parenteral antibiotics in the IQVIA MIDAS data available to us. The IQVIA MIDAS data did not distinguish between antibiotic sales for children and adults. We estimated DDDs per 1,000 inhabitants per day (DIDs) per country, using annual population estimates from 2019 World Bank country population.^16^

### Definitions

Antibiotics were categorised as Access, Watch, or Reserve according to the 2023 WHO AWaRe classification.^9^ Our analysis considered only Access and Watch antibiotics as Reserve, not-recommended, and unclassified antibiotics are of very low volumes and predominantly used in the hospital setting for complex, difficult-to-treat infections.

We will use the term “outpatient & community sectors” from here on to refer to antibiotic use outside of the inpatient healthcare sector. Broadly, the outpatient & community sectors included ambulatory care in hospitals and clinics, primary care, local pharmacies, care homes, and other sectors.

We defined the “Retail sector” as all sectors that are outside of the hospital (that is, excluding both outpatient (e.g. in clinics) and inpatient antibiotic use in hospitals) in the IQVIA MIDAS data.

### Calculation procedure to estimate oral antibiotic use across different sector

The observed ratio of oral-to-parenteral antibiotics in the Global-PPS inpatient dataset was determined for each country. A Bayesian multilevel model, assuming random country-level intercepts, was used with hospital-level aggregated inpatient antibiotic use data to estimate the 95% credible interval (CrI) around the estimate of the oral-to-parenteral (Appendix text 1). In the base case scenario, we assumed that parenteral antibiotic formulations were consumed exclusively in hospital facilities (sensitivity analyses relaxing this assumption were performed and are described in the section below).

Subsequently, the observed ratio of oral-to-parenteral antibiotics per country was multiplied to the total parenteral antibiotic in the IQVIA MIDAS Sales data to estimate national oral antibiotic use in the outpatient & community sectors (Figure 1). Data analysis was performed using R software version 4.3.2.^17^

**Figure 1.**
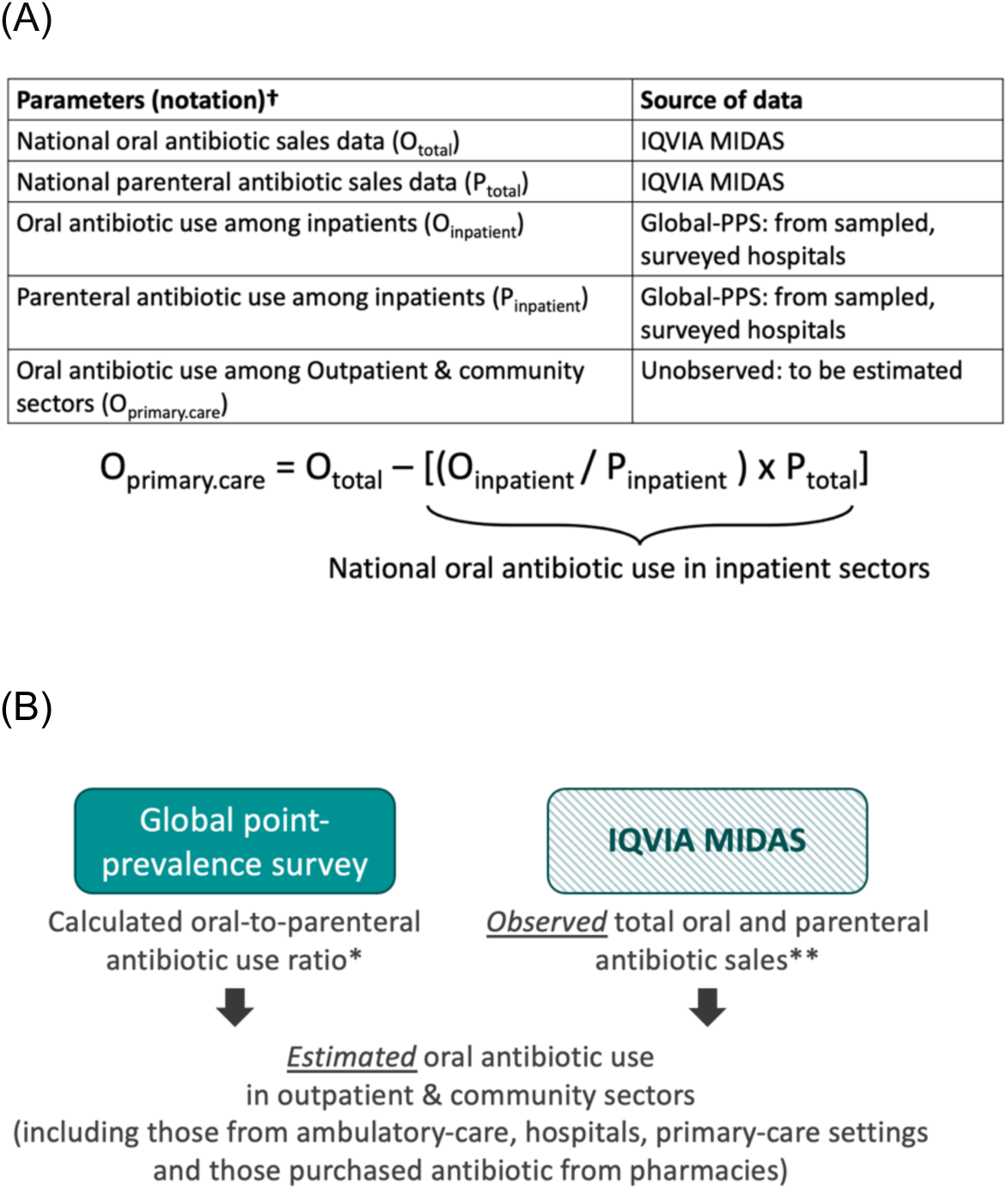
A conceptual diagram of calculation process (A) and data flow (B) to estimate antibiotic use across different sectors, and validate the estimates. **Footnote:** ^†^Defined Daily Dose (DDD) was used in the calculation to estimate the national oral antibiotic use in inpatient sector. *Global point-prevalence survey data representing inpatient antibiotic use in the survey hospitals of the eight countries under analysis in 2019 was used. **sales data was used by the authors to represent the overall country-level antibiotic use in the eight countries under analysis from the following source: IQVIA MIDAS Quarterly Sales data for 2019, reflecting estimates of real-world activity. Copyright IQVIA. All rights reserved.

### Validating the final estimates against antibiotic sales data from the retail sector

To validate the calculated estimates of national oral antibiotic use across inpatient, and outpatient & community sectors, we extracted data on oral antibiotic sales from the retail sector in the IQVIA MIDAS dataset. We then calculated the discrepancies between the estimated outpatient & community oral antibiotic use and the observed sales data from the retail sector. We expected to see smaller differences between the estimated national oral antibiotic and the observed oral antibiotic sales from the retail sector in the IQVIA MIDAS dataset in countries with relatively strong regulations on community antibiotic use, such as Belgium, the Netherlands, Canada, Singapore, and the United Kingdom, compared to countries with multiple alternative channels of antibiotic purchasing, including over-the-counter sales without a prescription.

### Sensitivity analyses

Additionally, we conducted two sensitivity analyses to assess the robustness of our findings to the calculated inpatient oral-to-parenteral ratios. In this set of sensitivity analyses, we assumed the oral-to-parenteral ratio was fifty percent lower and fifty percent higher, respectively, than observed in the Global-PPS 2019 data. Moreover, two sensitivity analyses to assess the robustness of the findings to the assumption that parenteral antibiotics were solely used in the inpatient sectors were performed. These two analyses assumed (1) 90% and (2) 50% of the national parenteral antibiotics sales were used in the inpatient sector.

## Results

### Antibiotic use in the eight countries

The total antibiotic sales in 2019 across the eight countries was 5.59 billion DDDs (9.1 DIDs) in the IQVIA MIDAS antibiotic data. Of those antibiotics, 65% (3.49 billion DDDs) were included in the 2023 WHO Essential Medicine List.^18^ The total Access and Watch antibiotic use was 5.36 billion DDDs (96% of total use). The observed total oral antibiotics use ranged from 4.95 DIDs in the Philippines to 24.27 DIDs in Belgium (Appendix Table 2).

**Table 1.** Observed inpatient antibiotic use from the 2019 Global Point Prevalence Survey study data of eight countries.

|  |  | **Observed absolute antibiotic use measured in number of DDD prescribed/100 surveyed adults on the day of the PPS** |  |  |
| --- | --- | --- | --- | --- |
| **Countries\*** | **AWaRe category** | **Oral antibiotics** | **Parenteral antibiotics** | **Estimates of oral-to-parenteral antibiotic ratios (95% credible interval)** |
| Belgium | Access | 8.3 | 18.4 | 0.49 (0.40-0.61) |
| Belgium | Watch | 5.6 | 10.6 | 0.55 (0.46-0.65) |
| Canada | Access | 8.0 | 12.8 | 0.77 (0.51-1.19) |
| Canada | Watch | 6.3 | 17.1 | 0.42 (0.30-0.58) |
| China | Access | 4.3 | 10.8 | 0.53 (0.25-1.09) |
| China | Watch | 2.4 | 58.3 | 0.05 (0.03-0.09) |
| Netherlands | Access | 9.3 | 22.1 | 0.47 (0.19-1.05) |
| Netherlands | Watch | 7.4 | 14.7 | 0.34 (0.15-0.79) |
| Philippines | Access | 9.0 | 12.5 | 0.58 (0.43-0.77) |
| Philippines | Watch | 25.9 | 30.3 | 0.84 (0.67-1.05) |
| Saudi Arabia | Access | 5.4 | 26.2 | 0.23 (0.12-0.49) |
| Saudi Arabia | Watch | 6.6 | 51.8 | 0.16 (0.10-0.27) |
| Singapore | Access | 13.0 | 18.5 | 0.62 (0.31-1.22) |
| Singapore | Watch | 7.5 | 16.5 | 0.41 (0.23-0.73) |
| United Kingdom | Access | 17.6 | 23.2 | 1.01 (0.56-1.80) |
| United Kingdom | Watch | 8.7 | 14.9 | 0.57 (0.37-0.89) |

**Table 2.**
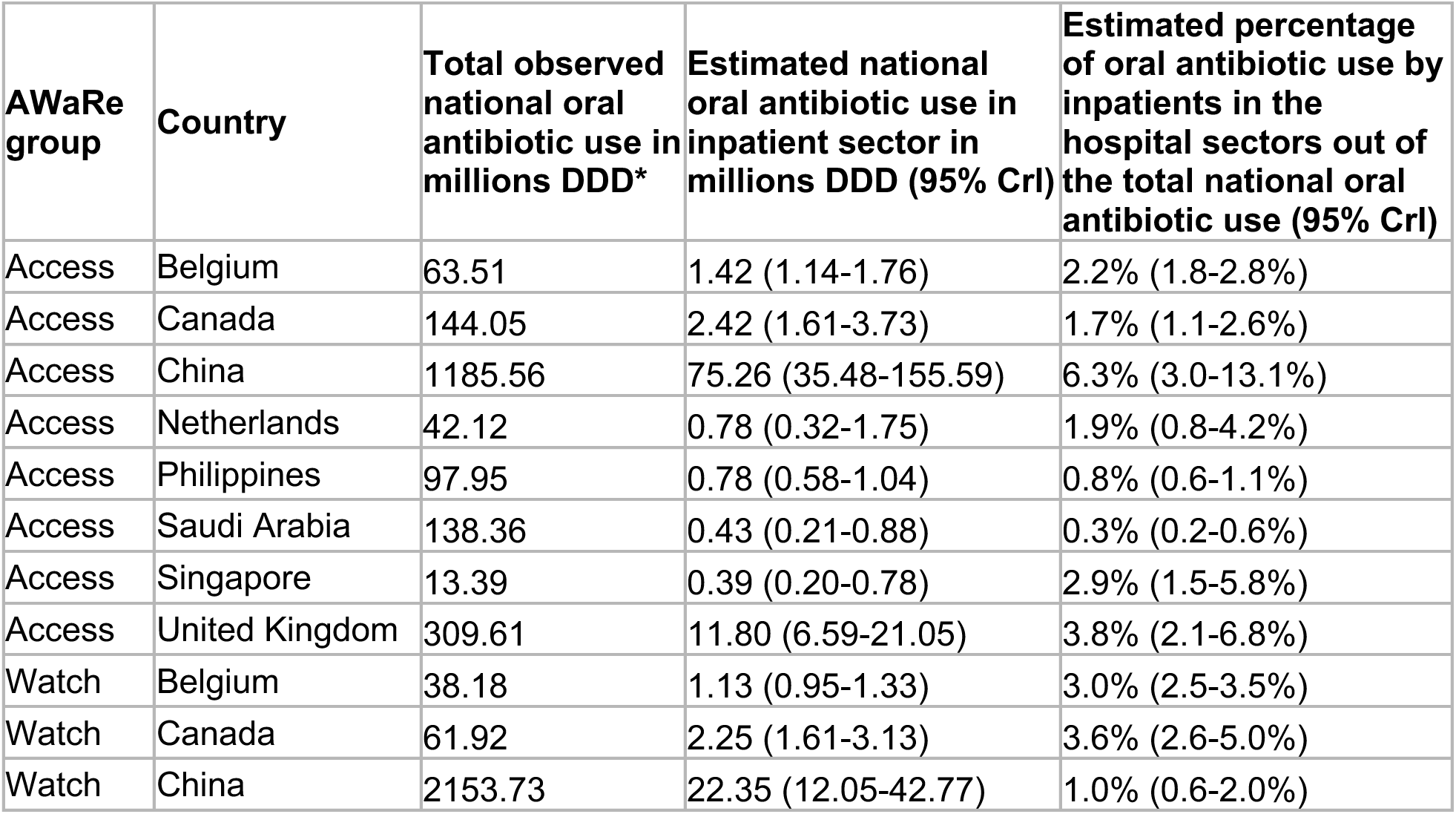
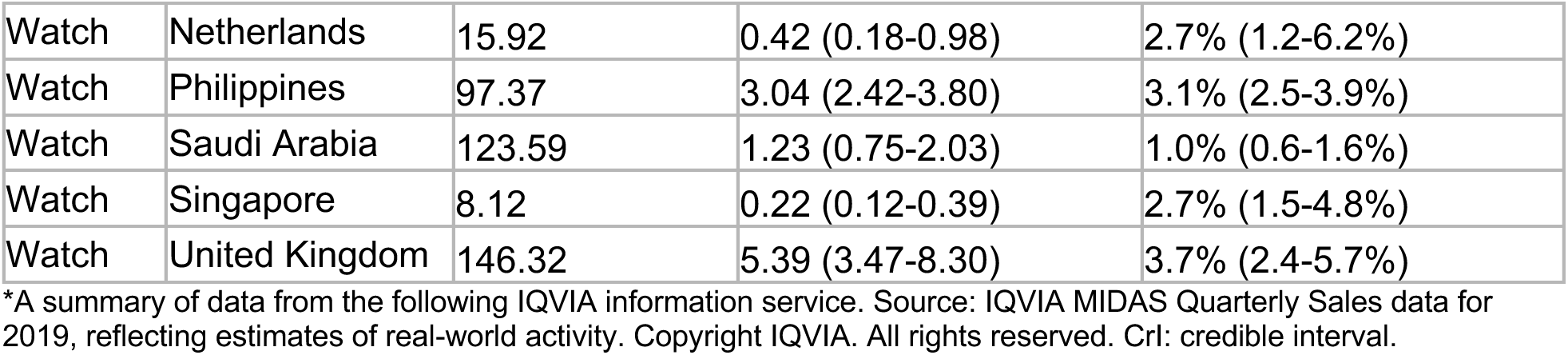
The overall observed national oral Access and Watch antibiotic use compared to the estimated oral antibiotic use within inpatient sectors.

The 2019 Global-PPS study data from the eight countries reported 16,351 DDDs (54.4 DDDs per 100 surveyed adult patients on the day of the PPS) of Access and Watch antibiotics (Table 1). There were variations in antibiotic choice across the eight countries (Appendix Figure 1). For example, amoxicillin/clavulanic acid ranked among the top three most commonly prescribed Access antibiotics in surveyed hospitals for inpatients in Belgium, the Netherlands, and Singapore, but not in the other five countries.

### Oral-to-parenteral antibiotic use ratio in Global-PPS data

Across all eight countries, the ratio of oral-to-parenteral antibiotic use in the Global-PPS data was below the value of 1, except for Access antibiotics in the United Kingdom (1.01 [95%CrI: 0.56-1.80]), implying oral and parenteral antibiotic usage was similar among the hospital inpatient population (Table 1 and Appendix Figure 2 & 3). The ratios of oral-to-parenteral antibiotic use were similar across three European countries. Small oral-to-parenteral ratios were observed for Watch antibiotics in China and for Access and Watch antibiotics in Saudi Arabia, suggesting there were large variation in the relative volumes of oral and parenteral antibiotics used in the hospital inpatient population.

### Estimated proportion of oral antibiotic usage in inpatient settings

After applying the ratios derived from the Global-PPS data to the IQVIA MIDAS data, estimates indicated that the proportion of oral antibiotics were used by inpatients ranged from 0.3% (95%CrI: 0.2-0.6%) to 6.3% (95%CrI: 3.0-13.1%) across the eight countries analysed (Table 2; Appendix table 3 and 4). When testing different oral-to-parenteral ratios, even with a ratio 1.5 times the observed value, over 94% of oral antibiotics were still used in non-inpatient sectors for most countries, except China. Similarly, when examining the assumption that parenteral antibiotics were predominantly used in inpatient settings, even if 50% would be used in non-inpatient settings, the estimated proportion of parenteral Access and Watch antibiotics used in inpatients remained above 90% in most of the countries included in the analysis (Appendix table 4).

**Table 3.** Variation between the estimated national oral antibiotic use among primary health care and observed oral antibiotics purchased by retail sectors.

| AWaRe | Country | Estimated national oral antibiotic use in the primary health care sectors in millions DDD (95% CrI) | Observed oral antibiotics purchased by retail sector in the IQVIA MIDAS 2019 data in millions DDD | Absolute difference in millions DDD* | Absolute difference in DID** |
| --- | --- | --- | --- | --- | --- |
| Access | Belgium | 62.10 (61.75-62.38) | 60.352 | 1.744 | 0.416 |
| Access | Canada | 141.63 (140.32-142.44) | 138.748 | 2.880 | 0.210 |
| Access | China | 1110.30 (1029.97-1150.08) | 767.836 | 342.460 | 0.666 |
| Access | Netherlands | 41.34 (40.36-41.79) | 40.418 | 0.918 | 0.145 |
| Access | Philippines | 97.17 (96.91-97.37) | 94.698 | 2.474 | 0.063 |

| **AWaRe** | **Country** | **Estimated national oral antibiotic use in the primary health care sectors in millions DDD (95% CrI)** | **Observed oral antibiotics purchased by retail sector in the IQVIA MIDAS 2019 data in millions DDD** | **Absolute difference in millions DDD\*** | **Absolute difference in DID\*\*** |
| --- | --- | --- | --- | --- | --- |
| Access | Saudi Arabia | 137.94 (137.48-138.15) | 110.401 | 27.537 | 2.202 |
| Access | Singapore | 13.00 (12.62-13.19) | 13.014 | -0.015 | -0.007 |
| Access | United Kingdom | 297.81 (288.56-303.02) | 256.210 | 41.602 | 1.705 |
| Watch | Belgium | 37.05 (36.85-37.23) | 32.671 | 4.382 | 1.045 |
| Watch | Canada | 59.67 (58.79-60.31) | 58.146 | 1.525 | 0.111 |
| Watch | China | 2131.38 (2110.97-2141.68) | 1127.783 | 1003.597 | 1.953 |
| Watch | Netherlands | 15.50 (14.94-15.74) | 15.135 | 0.364 | 0.057 |
| Watch | Philippines | 94.32 (93.56-94.94) | 91.609 | 2.713 | 0.069 |
| Watch | Saudi Arabia | 122.36 (121.56-122.84) | 91.679 | 30.683 | 2.453 |
| Watch | Singapore | 7.91 (7.73-8.00) | 8.102 | -0.195 | -0.094 |
| Watch | United Kingdom | 140.93 (138.02-142.85) | 128.733 | 12.199 | 0.500 |

\*Absolute differences between point estimate of national oral antibiotic use in the community & primary health sectors and the observed oral antibiotic purchased by retail sectors. \*\*Absolute difference in DID is the absolute difference in DDD standardised by the country population size; and in 2019 the populations size of Belgium, Canada, China, Netherlands, Philippines, Saudi Arabia, Singapore, and United Kingdom were 11,488,980; 37,601,230; 1,407,745,000; 17,344,874; 108,116,622; 34,268,529; 5,703,569; and 66,836,327, respectively. DDD: defined daily dose; DID: DDDs per 1000 inhabitants per day. Source: Based on data from the following IQVIA information service: IQVIA MIDAS Quarterly Sales data for 2019, reflecting estimates of real-world activity. Copyright IQVIA. All rights reserved

### Differences between estimated oral antibiotics and observed oral antibiotics in retail sectors

We used the data on oral antibiotic sales from the retail sector in the IQVIA MIDAS dataset to validate the estimated oral antibiotic use across inpatient, and outpatient & community sectors. The estimated oral antibiotic use outside of the inpatient healthcare sectors was larger than the observed oral antibiotics from the retail sector (all sectors that are outside of the hospital that is, excluding both outpatient [e.g. in clinics] and inpatient antibiotic use in hospitals) in all countries except for Singapore (Table 3). This could be due to multiple channels of antibiotic access, including retail sectors and outpatient care in hospitals and/or overstock within the community and primary health care sector. The differences between the estimated oral antibiotic use of outpatient & community sectors and the observed oral antibiotic purchased by the retail sector were less than 2 DID, except in Saudi Arabia. This confirmed the expectation that most of the primary health care antibiotics were purchased through the retail sectors in high-income settings where community antibiotic use is generally well-regulated. In all high-income settings, the estimated oral Access antibiotic use was higher than the estimated oral Watch antibiotics in the primary health care sectors. In China and the Philippines, higher oral Watch antibiotic was observed.

## Discussion

This study demonstrates the potential of using antibiotic administration routes as a proxy to estimate antibiotic use across different healthcare sectors. We estimate that less than 7% of national oral Access and Watch antibiotics procurement was used in the inpatient sector. Our finding supports the expectation that the great majority of oral antibiotics are used outside of the inpatient sector in the selected countries.^19–21^ We observed variations in oral-to-parenteral use ratios among inpatients across the eight participating hospitals in the Global-PPS 2019 data. Amongst the Access antibiotics, the oral-to-parenteral ratio ranged from 0.23 in Saudi Arabia, suggesting substantially lower oral compared to parenteral antibiotic use, to 1.01 in the UK, suggesting similar oral and parenteral Access antibiotic use. These variations are likely due to differences in antibiotic use patterns between countries. In the United Kingdom, oral amoxicillin (-/+ clavulanic acid) was the most widely used antibiotic (Appendix Figure 1) in the Global-PPS 2019 data, and its use was higher than the parenteral preparation of the same drug. It is not possible to comprehensively assess the reasons for higher oral antibiotic use compared to parenteral use without longitudinal antibiotic use and clinical data. However, one of the plausible reasons would be the implementation of the UK National Institute for Health and Care Excellence (NICE) antimicrobial stewardship guidelines on switching from intravenous to oral antibiotics.^22^

Rapid early parenteral to oral switching accompanied by a high hospital discharge rate could contribute to the observed high oral antibiotic use.^23^ Our observed variations in antibiotic use patterns across different countries could also be due to differences in healthcare provision, antimicrobial stewardship programmes, differences in infectious disease epidemiology,^24^ causative pathogen burdens in hospitals across different settings^1^, availability of parenteral and oral formulations of different antibiotics, and clinical characteristics of patients. This further highlights the importance of collecting local data to support evidence-based guidelines for targeted antimicrobial stewardship programmes.

We observed less variation in oral-to-parenteral antibiotic use ratios in the Watch antibiotics. All countries had ratios less than 1 suggesting higher parenteral compared to oral Watch antibiotic use. Watch antibiotics are often used for patients with severe infections who may need high dose intravenous antibiotics or may not tolerate oral medicines. Nevertheless, overuse of parenteral Watch antibiotics is also likely in many settings.^25^ Moreover, some Watch antibiotics such as ceftriaxone, commonly used for suspected sepsis, only have parenteral formulations and were among the top 90% of Watch antibiotics used in most hospitals in the Global-PPS 2019 (Appendix Figure 1). The oral-to-parenteral ratio of Watch antibiotics in the Philippines was large, and the upper bound of the uncertainty interval was above 1 (oral-to-parenteral ratio: 0.84 [95%CrI: 0.67-1.05]). One reason for this observation is the high oral cefuroxime use for surgical prophylaxis in the 31 participating hospitals in 2019. In the Philippines, a high rate of surgical prophylaxis prescriptions was noted, especially in Obstetrics and Gynaecology departments. Previous studies have shown low compliance with guidelines on antibiotic choice, route, and duration in surgery, with prolonged use being common.^26,27^

The estimated low proportion of national oral Access and Watch antibiotics that were used in the inpatient sectors was robust against a wide range of calculated inpatient oral-to-parenteral antibiotic ratio values (Appendix table 3). For instance, even when assuming a high inpatient oral-to-parenteral ratio of 1.5 times the observed ratio, most countries would still be expected to have around 95% of national oral antibiotics used in the non-inpatient sectors. One exception was China, which had a low overall proportion of oral antibiotic procurement (Appendix Table 2). This aligns with a previous study from China, which reported oral antibiotics accounted for 64% of national antibiotic use.^28^ Nevertheless, the overall observed oral (6.5 DID) and parenteral (1.4 DID) antibiotic sales in IQVIA MIDAS 2019 data was lower than reported by Yang et al (oral and parenteral antibiotic usage was 9.30 DID and 5.15 DID, respectively) from data of public health institution and during COVID-19.^28^

Overall, the findings from our study are consistent with the WHO GLASS-AMU’s suggestion that oral antibiotic use can be a surrogate for use in community and primary care.^7^ However, more granular data on antibiotic use from different health sectors are needed to inform targeted antimicrobial stewardship programmes. To achieve the target of at least 70% of human antibiotic use being Access antibiotics,^3,29^ our findings suggest that reducing national and global oral Watch antibiotic use would be essential. The volume of Watch oral antibiotic procurement was higher than Access oral antibiotic procurement in China, the Philippines, and Saudi Arabia (Table 2), and the estimated percentage of Watch antibiotic usage by the inpatient sector was low.

There are several key limitations to this study. First, the main analysis assumes all parenteral antibiotic use occurs in inpatient settings. While this assumption holds in some settings such as Lebanon, where 99% of antibiotic use in the community is represented by oral formulations,^30^ outpatient parenteral antibiotic use exists in the United States (since 1974)^31^ and other countries (including Australia, New Zealand, Canada, Singapore, and the United Kingdom).^32–34^ Our sensitivity analyses suggest that even in settings where 50% of parenteral antibiotics are used in non-inpatient sectors, the estimated proportion of parenteral Access and Watch antibiotics used in inpatients would still remain above 90% in most of the countries included in the analysis (Appendix Table 4). However, future studies with detailed hospital antibiotic use data would be needed to systematically assess the ratio of oral-to-parenteral antibiotic use in hospitals of different health systems, particularly with varying levels of Out Patient Antibiotic Use (OPAT) services. Second, our analysis only included two low- and middle-income countries, limiting generalisability to others. In settings such as China, with a high overall proportion of parenteral antibiotic use, the proportion of Access antibiotics used in outpatient & community sectors would be lower than 95% (Table 2). Third, our study assumes a stable hospital inpatient oral-to-parenteral ratio over time. This assumption may not be true in certain settings, for example due to seasonal infectious disease burdens. Moreover, the coverage of hospitals participating in the Global-PPS 2019 study varies across the eight countries included in the analysis (Appendix Table 1), which affects the generalisability of the estimated oral-to-parenteral ratios for the country-level inpatient population. For instance, data from only two hospitals in the Netherlands—one with 188 adult ward beds and the other with 474 beds—were included in the estimation of oral-to-parenteral ratios. This limitation highlights the importance of a future study to explore efficient strategies for a representative sampling frame for antibiotic use point-prevalence surveys across both primary care and hospital settings. Moreover, in this proof-of-concept study, we only used data from the adult inpatient samples of the Global-PPS 2019 study to estimate the oral-to-parenteral antibiotic use ratio, which does not sufficiently represent the paediatric antibiotic use pattern. This ratio was applied to the IQVIA MIDAS data on the overall national-level antibiotic procurement for the whole population including adults and paediatrics given the IQVIA MIDAS sales data does not allow us to differentiate between paediatric and adult usage. However, it is expected that adult use constitutes the majority. Nonetheless, standardising the absolute measurement of antibiotic use in paediatric population is crucial to quantify and monitor national and global antibiotic use. Furthermore, our analysis does not reflect the volume of appropriate antibiotic use and may not capture antibiotic use purchased from informal or unlicensed sources. Finally, we observed variations in the oral-to-parenteral antibiotic use ratio across countries. Therefore, directly applying these ratios from one country to another should be avoided. Future analyses to validate the application of our approach to estimate national antibiotic use across healthcare sectors for settings with different antibiotic market structure and healthcare-seeking behaviours would be important.

## Conclusion

Our results suggest that where patient-level data are unavailable, alternative sources, such as antibiotic import data which includes routes of administration, can be used to estimate sector-specific national use. However, variations in antibiotic use patterns across different countries are large and innovative studies to collect antibiotic use efficiently data across different healthcare sectors are needed to support the development of locally tailored policies and interventions, and to validate our results. For instance, patterns of antibiotic use identified from macro-level import and other data could be validated using novel primary-care point-prevalence survey methods (Cook A et al. 2025^35^ and Boven A et al. 2025^36^). Sustainable citizen-led initiatives to collect community antibiotic use data could also play an important role in filling data gaps. Representative sampling frames for both surveys could then be combined with other data sets to provide robust estimates of population-level patterns of use to inform national antibiotic policies.

## Disclaimer

The findings and conclusions in this report are those of the authors and do not necessarily represent the official position of WHO or any of the institutions mentioned.

## Data Availability

The raw datasets used in this study are not publicly available.

## Acknowledgement

We would like to thank Peter Stephens for his valuable feedback on the draft of this manuscript and for the IQVIA MIDAS^®^ Quarterly Sales data for 2019 used in this article, which were obtained under license from IQVIA and reflect estimates of real world activity. Copyright IQVIA. All rights reserved. The statements, findings, conclusions, views, and opinions contained and expressed herein are not necessarily those of IQVIA.

We would like to thank the members of the Global-PPS network in the participating countries:

Erica Sermijn (ASZ Aalst, Aalst, Belgium), Katia Verhamme (OLV Hospital, Aalst, Belgium), Bruno Van Herendael (GZA Hospitals, Antwerp, Belgium), Helena Mertes (Ziekenhuis Netwerk Antwerpen (ZNA), Antwerp, Belgium), Ingrid Wybo (Universitair Ziekenhuis Brussel, Jette, Belgium), Johan Frans (Imelda Hospital, Bonheiden, Belgium), Geert Vanheule (General Hospital Rivierenland, Bornem, Belgium), Nathalie Gillard (Clinique Saint-Luc, Bouge, Belgium), Pauline Papin (Clinique Saint-Jean, Brussels, Belgium), Laetitia Brassinne (Europe Hospitals, Brussels, Belgium), Marc Vekemans (IRIS South Hospitals, Brussels, Belgium), Deborah Konopnicki (Saint-Pierre University Hospital, Brussels, Belgium), Sandrine Milas (CHU Charleroi, Charleroi, Belgium), Xavier Holemans (Grand Hôpital de Charleroi (GHdC), Charleroi, Belgium), Hilde Jansens (University Hospital of Antwerp, Edegem, Belgium), Sofie Bartholomeus (Sint-Dimpna Ziekenhuis, Geel, Belgium), Louis Ide (AZ Jan Palfijn Gent, Ghent, Belgium), Sophia Steyaert (AZ Maria Middelares , Ghent, Belgium), Anne-Marie Van den Abeele (General Hospital St. Lucas , Ghent, Belgium), Patricia Schatt (Clinique Notre Dame de Grâce, Gosselies, Belgium), Reinoud Cartuyvels (Jessa Ziekenhuis, Hasselt, Belgium), Danielle Van der beek (AZ Herentals, Herentals, Belgium), Nele Schrooten (Sint-Franciscusziekenhuis, Heusden-Zolder, Belgium), Aline Honoré (Centre Hospitalier Régional de Huy, Huy , Belgium), Lorenz Vanneste (AZ Groeninge, Kortrijk, Belgium), Eric Firre (Hopital de la Citadelle de Liege, Liège, Belgium), Christelle Vercheval (University hospital of Liège, Liège, Belgium), Wim Laffut (Heilig Hart Ziekenhuis, Lier, Belgium), Marc Vandevelde (Ziekenhuis Maas en Kempen, Maaseik, Belgium), Clara Ceyssens (AZ Voorkempen, Malle, Belgium), Peter Verbeeck (Heilig Hartziekenhuis, Mol, Belgium), Camelia Rossi (CHU Ambroise Paré, Mons, Belgium), Clotilde Visée (Hospital of Mons / Warquignies (Pôle hospitalier Jolimont), Mons, Belgium), Francois Kidd (CHU UCL Namur , Namur, Belgium), Ilisei Dragos (ASBL Jolimont, Hôpital Nivelles , Nivelles , Belgium), Bart Glibert (AZ Glorieux, Ronse, Belgium), Kay Versonnen (AZ Rivierenland, Rumst, Belgium), Frederik Van Hoecke (Sint Andries Hospital, Tielt, Belgium), Reinhilde Reybrouck (Regionaal Ziekenhuis H. Hart Tienen, Tienen, Belgium), Dana Van Kerkhoven (AZ Turnhout, Turnhout, Belgium), Sofie Colman (OLV van Lourdes Hospital, Waregem, Belgium), Benedicte Delaere (CHU UCL Namur, Yvoir, Belgium), Michiel Costers (Sint-Elisabeth Ziekenhuis, Zottegem, Belgium), Dominik Mertz (Hamilton Health Sciences, Hamilton, Ontario, Canada), Susan McKenna (Kingston Health Sciences Centre, Kingston, Ontario, Canada), Yu-Chen Lin (Vancouver Coastal Health, Lions Gate Hospital, North Vancouver, British Columbia, Canada), Anaïs Lauzon-Laurin (CHDL, CISSS de Lanaudière, Saint-Charles-Borromée, Québec, Canada), Louis Valiquette (CIUSSS de l’Estrie - CHUS, Sherbrooke, Québec, Canada), Jacqueline Roberts (Perth and Smiths Falls District Hospital, Smiths Falls, Ontario, Canada), Kevin Afra (Fraser Health, Surrey, British Columbia, Canada), Yukun Chen (The 1st affiliated hospital of Jilin University, Changchun, China), Wenxiang Huang (The 1st affiliated hospital of Chongqing Medical University, Chongqing, China), Jingping Zhang (The First Hospital of China Medical University, Shenyang, China), Feng Changwen (The First People’s Hospital of Zhaoqing, Zhaoqing, China), Michiel van Rijn (Ikazia Ziekenhuis , Rotterdam, Netherlands), Valentijn Schweitzer (University Medical Center Utrecht, Utrecht, Netherlands), Therese Anne M. Suñe – Lagdameo (Corazon Locsin Montelibano Memorial Regional Hospital, Bacolod City, The Philippines),Melody C. Gulian (Saint Louis University Sacred Heart Medical Center, Baguio City, The Philippines), Johnson D. Palaypay (Bataan General Hospital, Balanga, The Philippines), Isolde Mayo (Cotabato Regional and Medical Center, Cotabato City, The Philippines), Rolando Jr. S. Padilla (Iloilo Doctor’s Hospital, Iloilo city, The Philippines), Ruby Ramos (Our Lady of Pillar Medical Center, Imus, The Philippines), Ma. Cecilia T. delos Reyes (Las Piñas General Hospital and Satellite Trauma Center, Las Pinas, The Philippines), Anna Lynda T. Bellen (Bicol Regional Hospital and Medical Center, Legazpi City, The Philippines), Bernadette T. Seludo (Victor R. Potenciano Medical Center (VRP Medical Center), Mandaluyong City, The Philippines), Lozel D. Villadore (Manila Doctors Hospital, Manila, The Philippines), Ethel Catherine M. Antonio (San Lazaro Hospital, Manila, The Philippines), Darel Gresan C. Quisil (Bukidnon Provincial Hospital - Maramag, Maramag, The Philippines), Angelica Martin (Asian Hospital Medical Center, Muntinlupa, The Philippines), Liezel C. Afaga (Ospital ng Muntinlupa, Muntinlupa, The Philippines), Mari Rose A. De Los Reyes (Research Institute for Tropical Medicine, Muntinlupa, The Philippines), Jemelyn U. Garcia (Research Institute for Tropical Medicine, Muntinlupa, The Philippines), Rhenalyn V. Bo (Research Institute for Tropical Medicine, Muntinlupa, The Philippines), Evelyn P. Hezeta (James L. Gordon Memorial Hospital, Olongapo, The Philippines), Marja B. Buensalido (The Medical City, Pasig City, The Philippines), Ma. Charmian Hufano (De Los Santos Medical Center, Quezon City, The Philippines), Raquel Victoria M. Ecarma (National Kidney and Transplant Institute, Quezon City, The Philippines), Jay Ron O. Padua (Philippine Children’s Medical Center, Quezon City, The Philippines), Cherie C. Araneta (Roxas Memorial Provincial Hospital, Roxas, The Philippines), Mary Shiela Ariola-Ramos (Cardinal Santos Medical Center, San Juan, The Philippines), Nori Jane G. Aroc (Southern Isabela Medical Center, Santiago, The Philippines), Susana N. Tizon (Eastern Visayas Regional Medical Center, Tacloban City, The Philippines), Nomi R. Aparece (St. Luke’s Medical Center-Global City, Taguig City, The Philippines), Mitzie Lou C. Osabel (Davao Regional Medical Center, Tagum City, The Philippines), Siok Ying Lee (Khoo Teck Puat Hospital, Singapore, Singapore), Hui Hiong Chen (National University Hospital, Singapore, Singapore), Sing Meng Choo (Ng Teng Fong General Hospital, Singapore, Singapore), Sock Hoon Tan (Tan Tock Seng Hospital, Singapore, Singapore), Fidelma Magee (Northern Health and Social Care Trust, Antrim, United Kingdom), Cairine Gormley (Altnagelvin Hospital and South West Acute Hospital, Western Heatlh and Social Care trust, Derry and Enniskillen , United Kingdom), Geraldine Conlon-Bingham (Southern Health and Social Care Trust, Portadown, United Kingdom)

## Funding

This work is part of ADILA (Antibiotic Data to Inform Local Action). ADILA is supported by the Wellcome Trust [222051/Z/20/Z] who had no role or responsibility in the study design, data collection, analysis, interpretation or writing the report.

BioMérieux is the sole private partner of the Global Point Prevalence Survey. BioMérieux had no role nor responsibility in the study design, data collection, data analysis, data interpretation, or writing the report, which was done under the sole responsibility of the University of Antwerp. Data are strictly confidential and stored anonymously at the coordinating centre of the University of Antwerp.

## Appendices

### Appendix text: Methods

#### Bayesian model with random intercept model

To estimate the uncertainty interval around the calculated oral-to-parenteral antibiotic use ratio, we fitted a Bayesian hierarchical model without any predictor variable. The oral-to-parenteral antibiotic use ratios calculated from data of each participating hospital were log-transformed to avoid taking the value 0. We define the model as follows. Let *Y_ij_* denote the oral-to-parenteral antibiotic use ratio on a log scale for hospital *i* in country *j*. The form of our model is

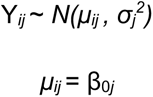

We assumed a vague prior for β that is normally distributed with mean 0 and a large variance (value of 5^2^) to represent a weakly informed prior. We assumed *σ* follows a Cauchy distribution with median 0 and scale parameter of 2.

In total, 248 oral-to-parenteral antibiotic use ratios were calculated from 125 participating hospitals in 2019 (Appendix Figure 2). For the participating hospitals without observations for a particular class of antibiotics (Access/Watch) observed on the day of the survey, we avoided assigning a value of 0 (which could result in an undefined mathematical value when calculating the ratio) by assuming that the use was minimal and assigning a value of 1 DDD. Of the 125 participating hospitals, there were 3 hospitals without an observed parenteral Access antibiotic, 2 without an observed parenteral Watch antibiotic, 2 without oral Access antibiotic, and 1 without observed oral Watch antibiotic on the day of the survey.

**Appendix table 1.** The number of hospitals participating in Global-PPS in 2019.

| Country | Number of hospitals participating in the 2019 survey | Median (minimum, maximum) number of beds in adult wards surveyed per participating hospital |
| --- | --- | --- |
| Belgium | 56 | 265 (30, 1024) |
| Canada | 14 | 277.5 (34, 641) |
| China | 4 | 127.5 (108, 142) |
| Netherlands | 2 | 331 (188, 474) |
| Philippines | 31 | 247 (12, 706) |
| Saudi Arabia | 6 | 124 (20, 283) |
| Singapore | 4 | 825 (668, 1393) |
| United Kingdom | 8 | 195.5 (48, 433) |

**Appendix table 2.** Observed combined Access and Watch antibiotic consumption from IQVIA MIDAS 2019 data of eight countries.

| Countries included in the analysis | *Observed oral antibiotic wholesales measure in millions DDD (DID) | *Observed parenteral antibiotic wholesales measure in millions DDD (DID) | Proportion of observed oral antibiotics |
| --- | --- | --- | --- |
| Belgium | 101,774,342 (24.27) | 5,010,667 (1.19) | 95.3% |
| Canada | 206,027,097 (15.01) | 8,847,642 (0.64) | 95.9% |
| China | 3,461,131,988 (6.74) | 708,353,513 (1.38) | 83.0% |
| Netherlands | 58,053,214 (9.17) | 2,915,291 (0.46) | 95.2% |
| Philippines | 195,423,103 (4.95) | 5,018,548 (0.13) | 97.5% |
| Saudi Arabia | 262,051,283 (20.95) | 9,682,879 (0.77) | 96.4% |
| Singapore | 21,599,786 (10.38) | 1,241,065 (0.60) | 94.6% |
| United Kingdom | 456,280,395 (18.70) | 22,127,364 (0.91) | 95.4% |

Footnote: DDD= defined daily dose; DID= DDD per 1,000 inhabitants. \*Based on data from the following IQVIA information service: IQVIA MIDAS Quarterly Sales data for 2019, reflecting estimates of real-world activity. Copyright IQVIA. All rights reserved.

**Appendix table 3.**
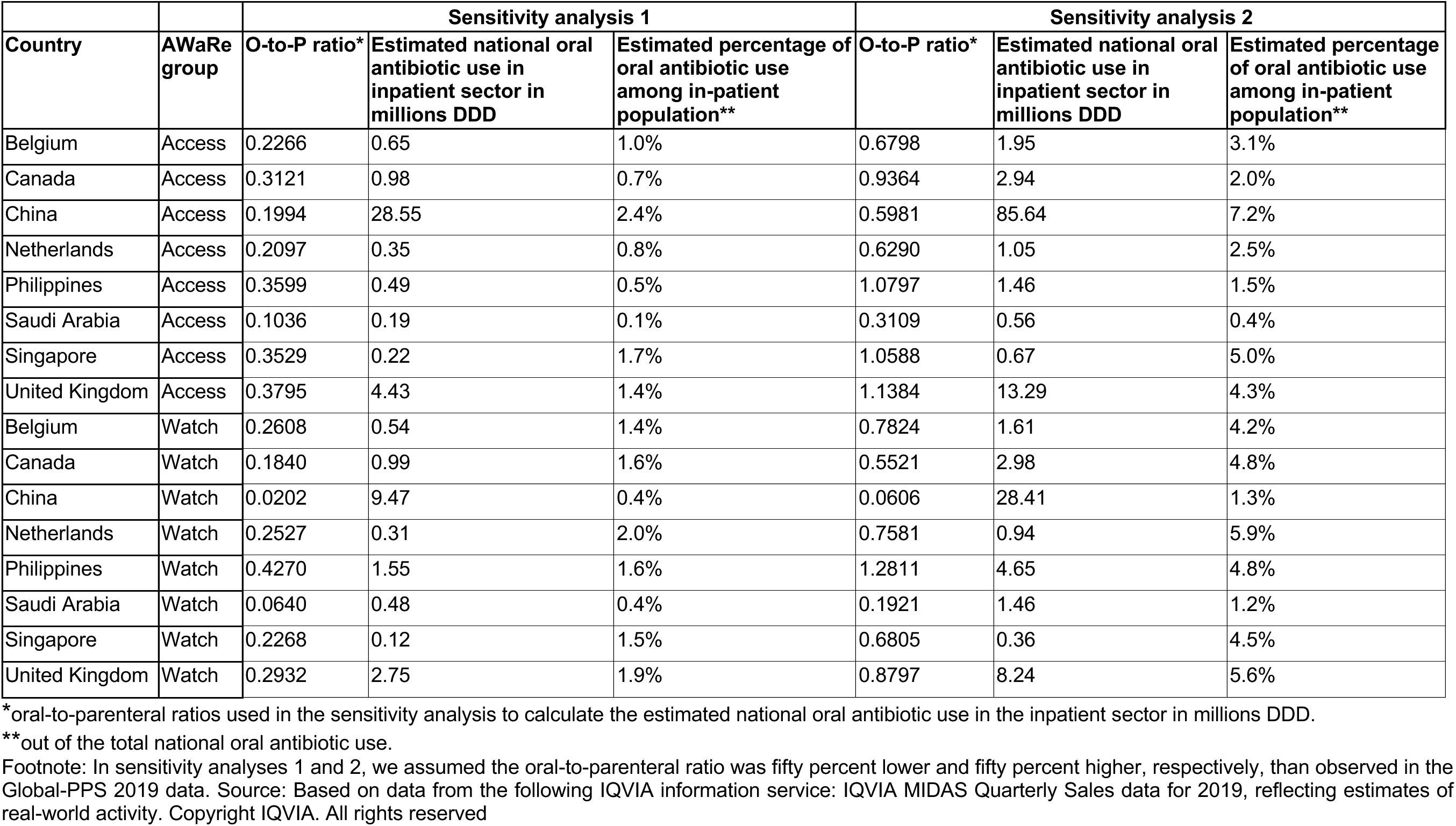
Results from the two sensitivity analyses to explore the robustness of the findings against the calculated oral-to-parenteral antibiotic use ratio.

**Appendix table 4.** Results from the two sensitivity analyses to explore the robustness of the findings against the assumption of all parenteral antibiotics being used in the inpatient sector.

|  |  | Sensitivity analysis 3 |  |  | Sensitivity analysis 4 |  |  |
| --- | --- | --- | --- | --- | --- | --- | --- |
| Country | AWaRe group | Proportion of parenteral antibiotics used in the inpatient sector | Estimated national oral antibiotic consumption in inpatient sector in millions DDD | Estimated percentage of oral antibiotic use among in-patient population** | Proportion of parenteral antibiotics used in the inpatient sector* | Estimated national oral antibiotic use in inpatient sector in millions DDD | Estimated percentage of oral antibiotic use among in-patient population** |
| Belgium | Access | 90% | 1.45 | 2.3% | 50% | 2.61 | 4.1% |
| Canada | Access | 90% | 1.82 | 1.3% | 50% | 3.27 | 2.3% |
| China | Access | 90% | 99.31 | 8.4% | 50% | 178.76 | 15.1% |
| Netherlands | Access | 90% | 0.68 | 1.6% | 50% | 1.23 | 2.9% |
| Philippines | Access | 90% | 0.60 | 0.6% | 50% | 1.08 | 1.1% |
| Saudi Arabia | Access | 90% | 0.08 | 0.1% | 50% | 0.15 | 0.1% |
| Singapore | Access | 90% | 0.30 | 2.2% | 50% | 0.53 | 4.0% |
| United Kingdom | Access | 90% | 6.55 | 2.1% | 50% | 11.80 | 3.8% |
| Belgium | Watch | 90% | 1.65 | 4.3% | 50% | 2.96 | 7.8% |
| Canada | Watch | 90% | 5.11 | 8.3% | 50% | 9.20 | 14.9% |
| China | Watch | 90% | 107.97 | 5.0% | 50% | 194.35 | 9.0% |
| Netherlands | Watch | 90% | 0.18 | 1.1% | 50% | 0.32 | 2.0% |
| Philippines | Watch | 90% | 2.84 | 2.9% | 50% | 5.12 | 5.3% |
| Saudi Arabia | Watch | 90% | 3.82 | 3.1% | 50% | 6.87 | 5.6% |
| Singapore | Watch | 90% | 0.45 | 5.5% | 50% | 0.81 | 10.0% |
| United Kingdom | Watch | 90% | 6.11 | 4.2% | 50% | 10.99 | 7.5% |

oral-to-parenteral ratio used in the sensitivity analysis to calculate the estimated national oral antibiotic use in inpatient sector in millions DDD
\*\*out of the total national oral antibiotic use.
Footnote: In sensitivity analyses 3 and 4, we assumed 90% and 50% of the national parenteral antibiotics procured were used in the inpatient sector. Source:
Based on data from the following IQVIA information service: IQVIA MIDAS Quarterly Sales data for 2019, reflecting estimates of real-world activity. Copyright IQVIA. All rights reserved

**Appendix Figure 1.**
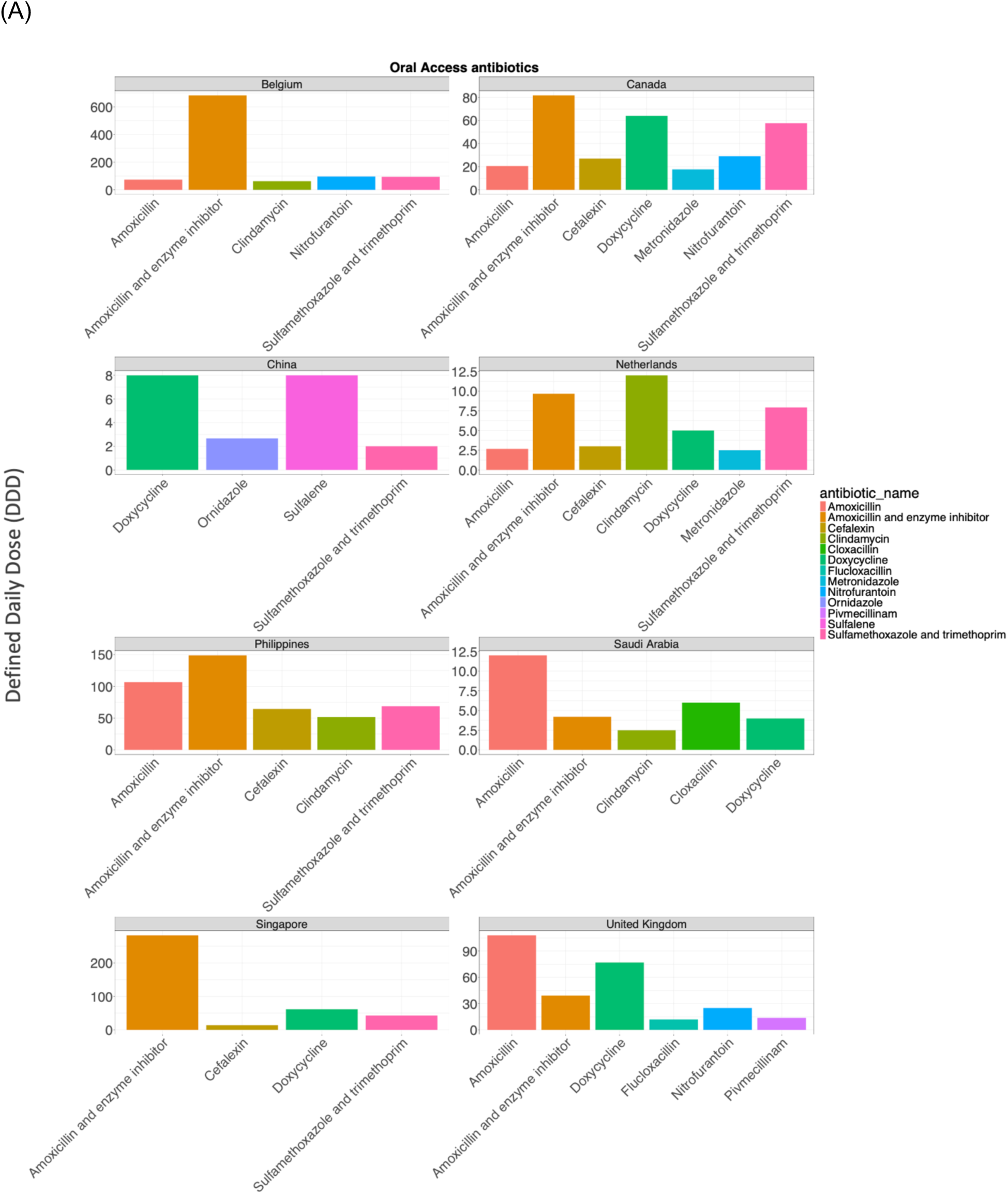
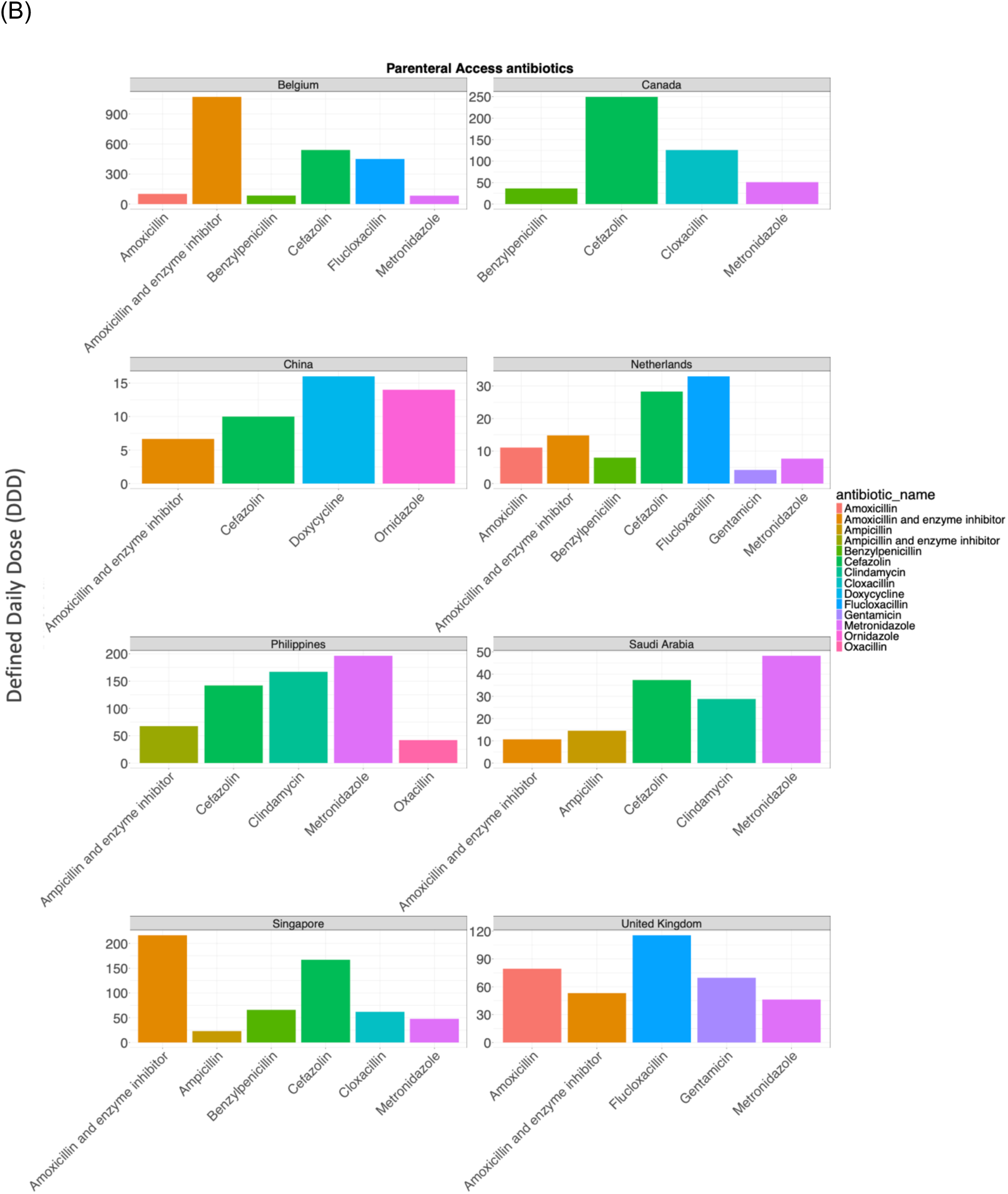
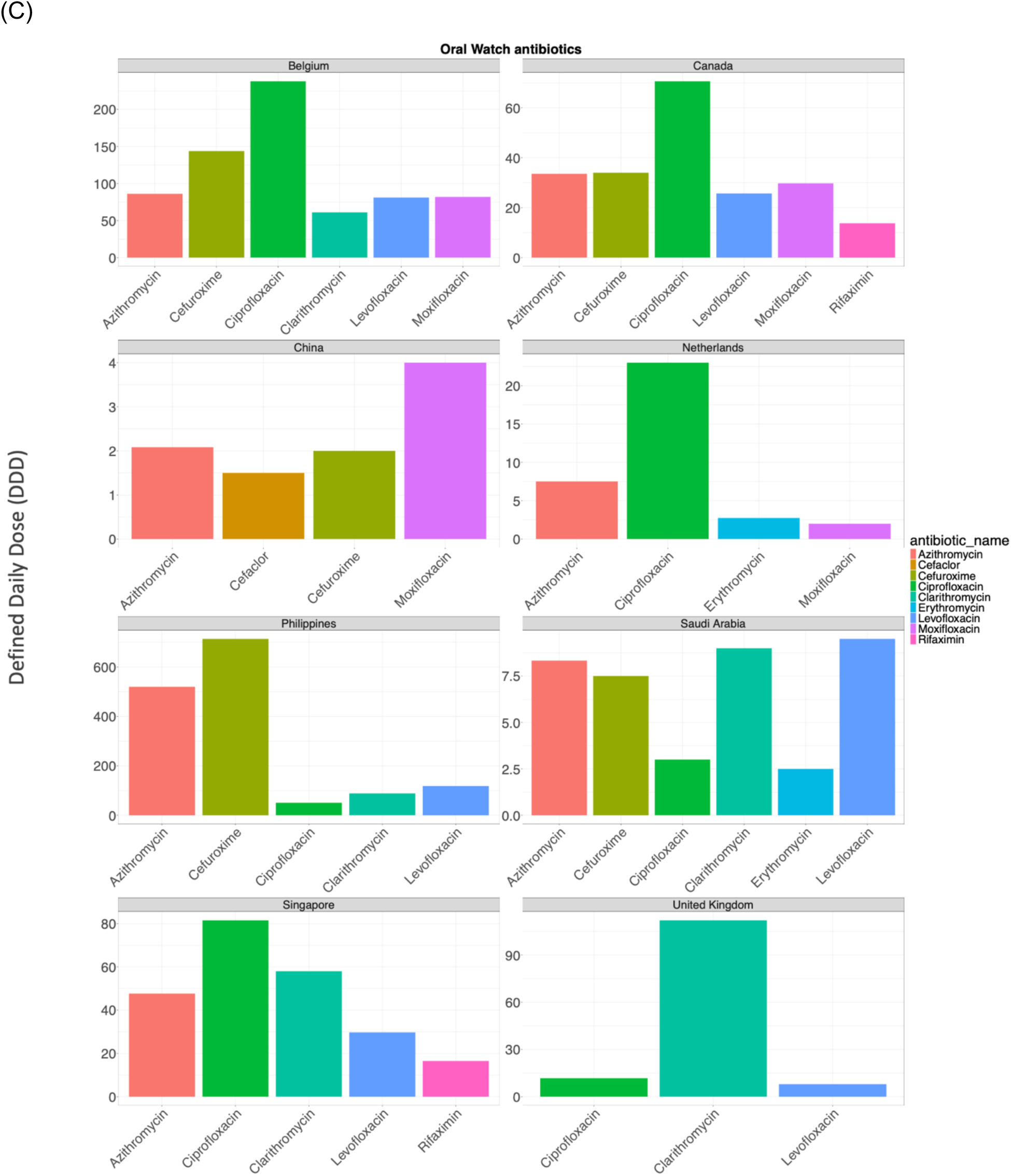
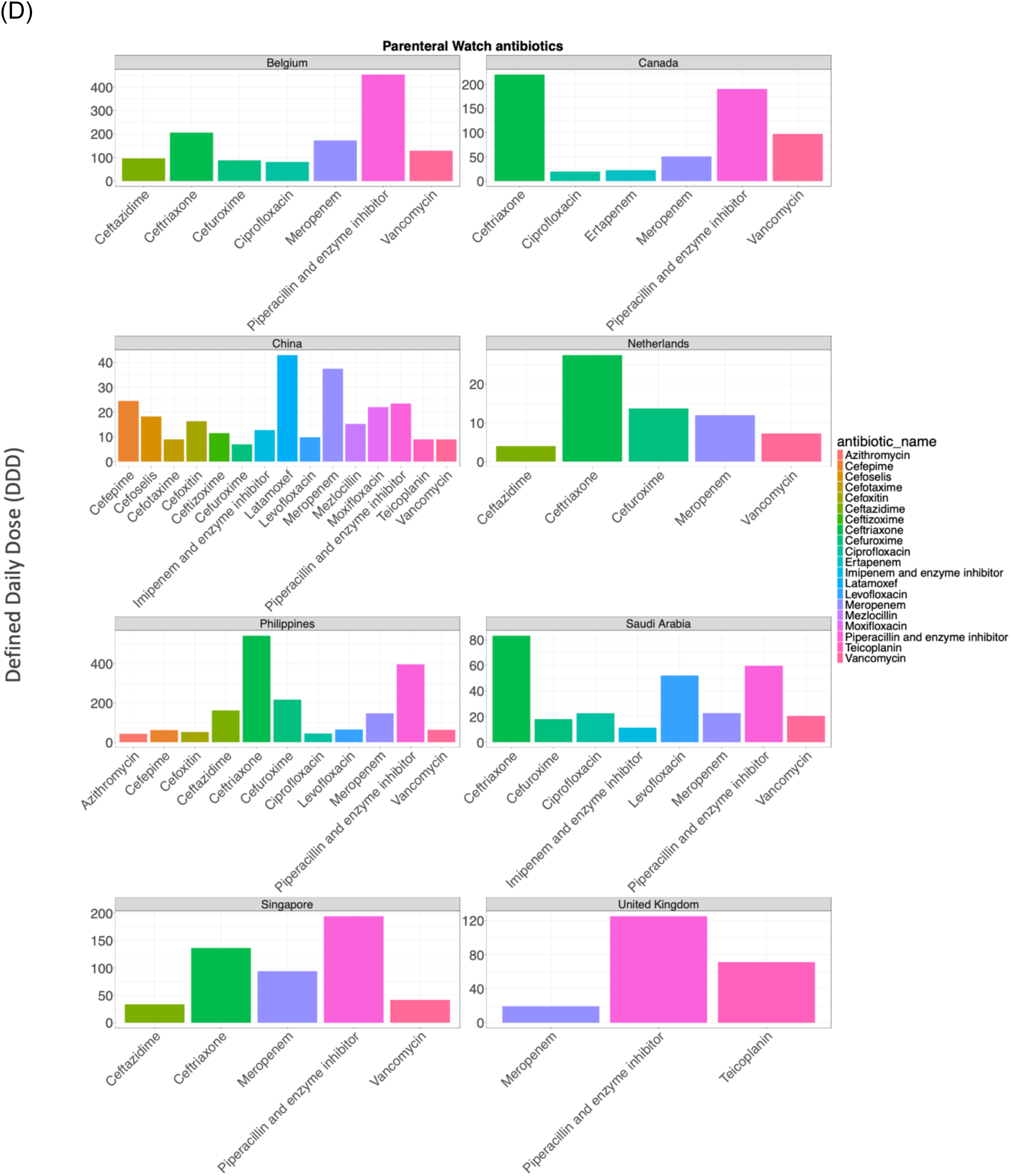
Distribution of different types of oral and parenteral antibiotics accounting for 90% of the prescriptions used by inpatients on the day that Global-PPS was performed in the participating hospital across the eight countries included in the analysis. The types of Access antibiotic (A and B) used varied by route of administration across the countries. For example, in China oral administration of amoxicillin or amoxicillin plus enzyme inhibitor was not reported in hospitals included in the 2019 Global-PPS survey, whereas the oral usage of both amoxicillin and amoxicillin plus enzyme inhibitor were the most widely used oral antibiotics in the other countries. Similarly, the number of different types of Watch antibiotics (C and D) varied across different countries and routes of administration. The graphs (A-D) were generated using the Global PPS 2019 data.

**Appendix Figure 2.**
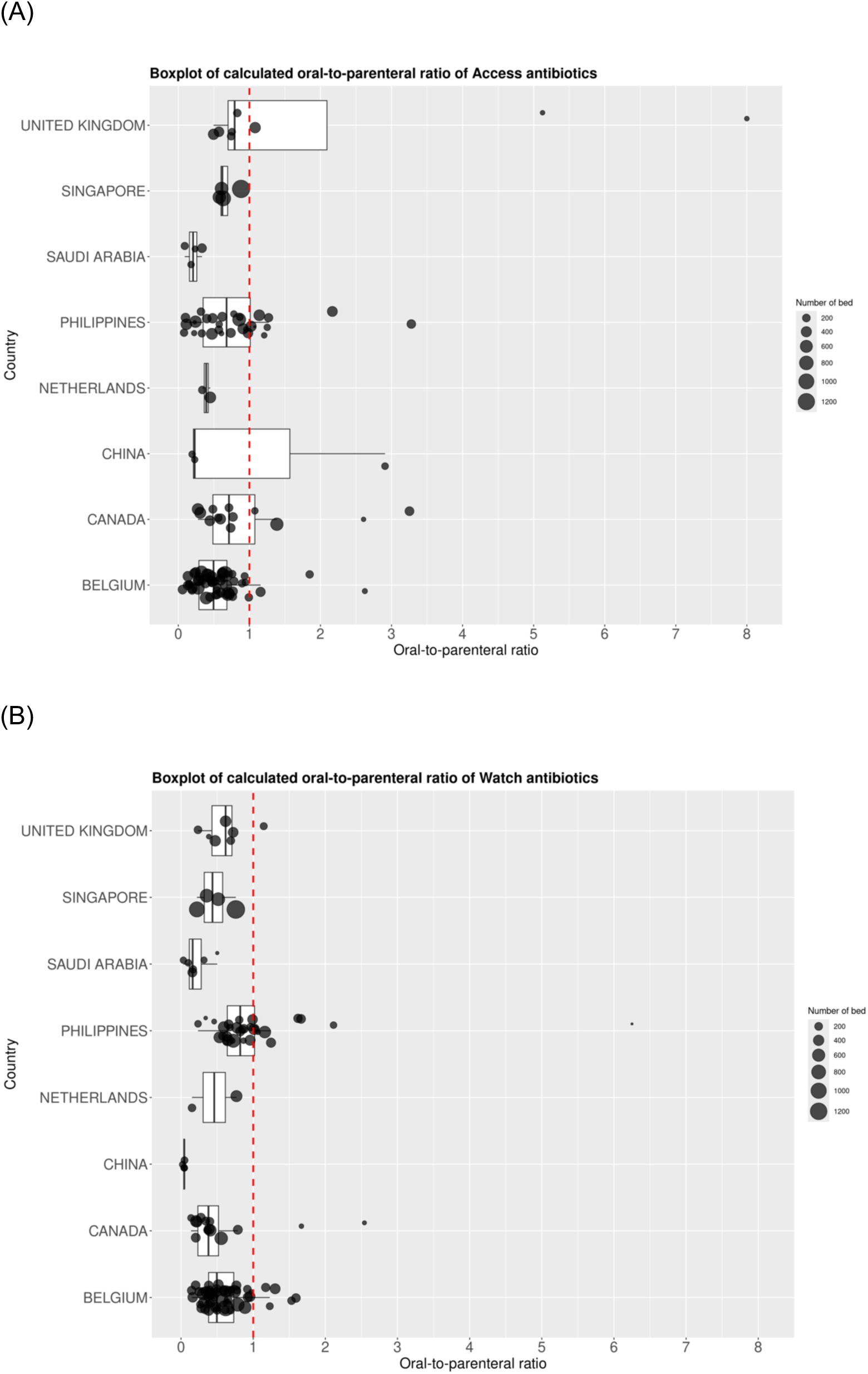
Distribution of oral-to-parenteral ratios across different surveyed hospitals by country, stratified by (A) Access and (B) Watch antibiotics.

**Appendix Figure 3.**
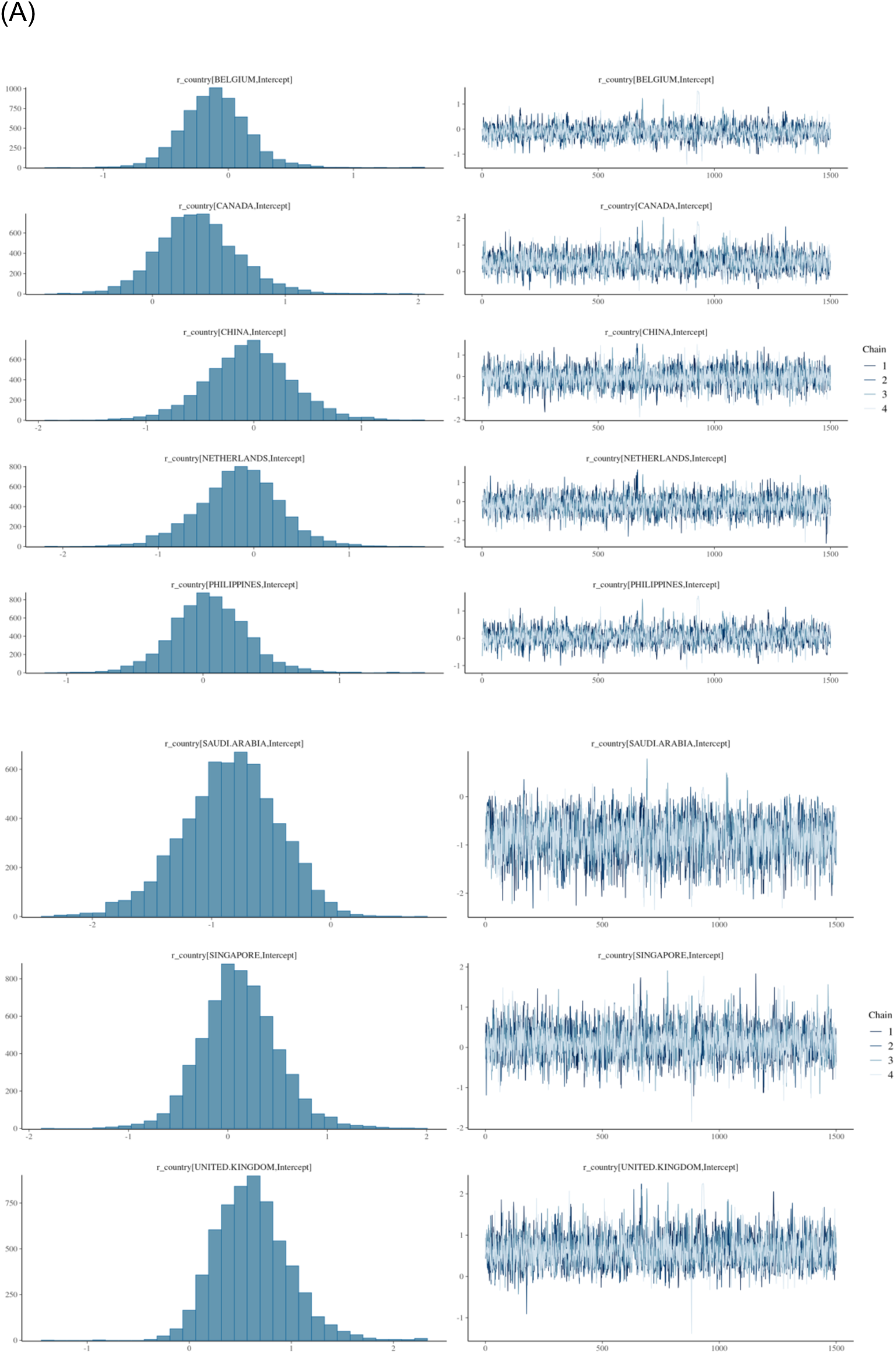
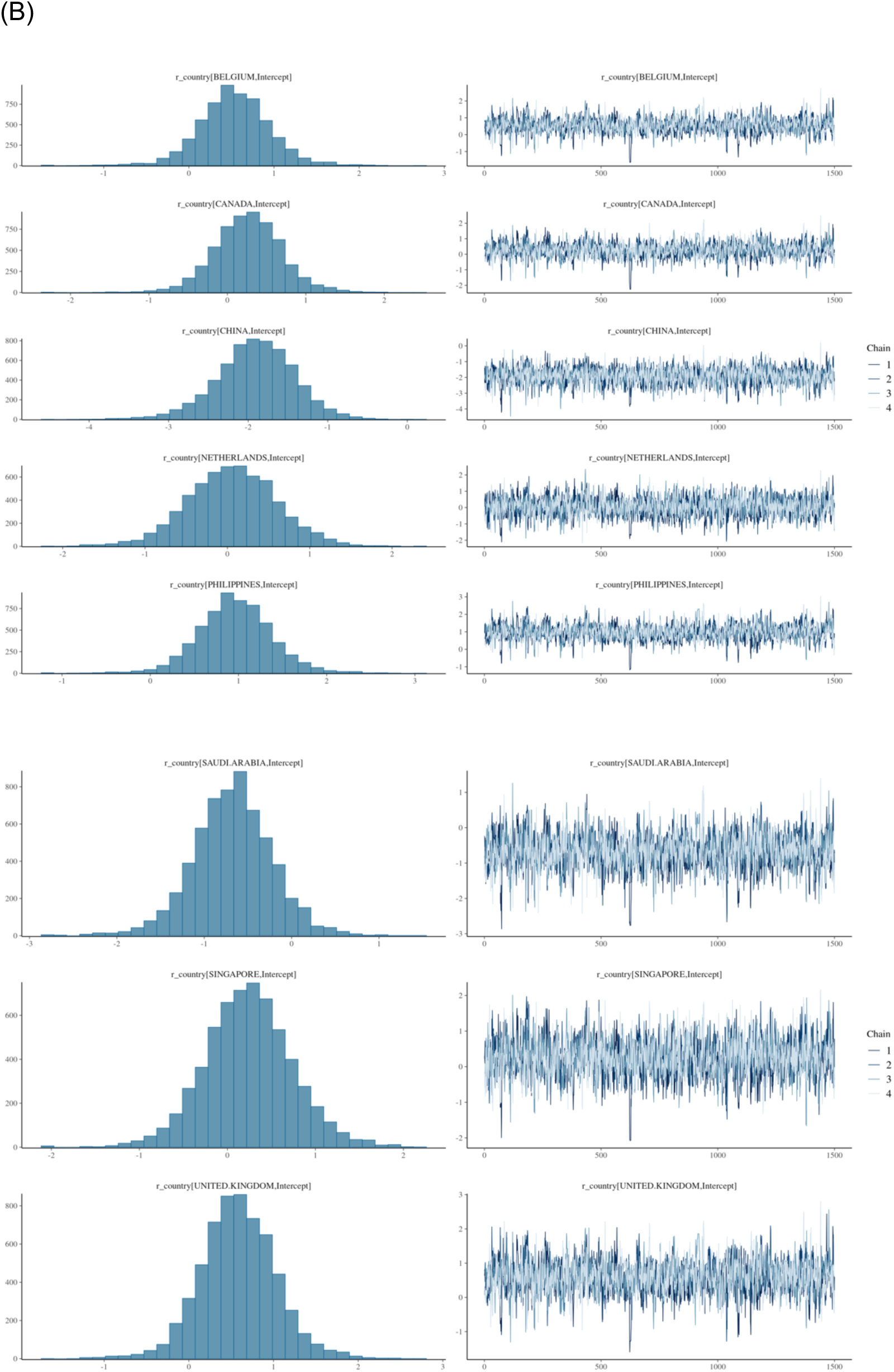
Posterior distributions from Bayesian random intercept models on log transformed oral-to-parenteral antibiotic use ratios by countries for (A) Access antibiotics and (B) Watch antibiotics.

